# Machine learning-based noise reduction for craniofacial bone segmentation in CT images

**DOI:** 10.1101/2022.06.26.22276925

**Authors:** Soh Nishimoto, Takuya Saito, Hisako Ishise, Toshihiro Fujiwara, Kenichiro Kawai, Maso Kakibuchi

**Affiliations:** Department of Plastic Surgery, Hyogo College of Medicine

**Keywords:** Computer-assisted tomography (CT), Cranio-facial bone, Metal artifact, U-net, DICOM

## Abstract

**Objective:** When high X-ray absorption rate materials such as metal prosthetics are in the field of CT scan, noise called metal artifacts might appear. In reconstructing a three-dimensional bone model from X-ray CT images, the metal artifacts remain. Often, the image of the scanning bed also remains. A machine learning-based system to reduce noises in the craniofacial CT images was constructed.

**Methods:** DICOM images of CT archives of patients with head and neck tumors were used. The metal artifacts and beds were removed from the threshold segmented images to obtain the target bony images. U-nets, respectively with the function loss of mean squared error, Dice and Jaccard, were trained by the datasets consisting of 5671 DICOM images and corresponding target images. DICOM images of 2000 validation datasets were given to the trained models and predicted images were obtained.

**Results:** The use of mean squared errors presented superiority to Dice or Jaccard loss. The mean prediction error pixels were 14.43, 778.57, and 757.60 respectively per 512 × 512 pixeled image.

**Discussion:** Dedicated to the delineation of craniofacial bones, the presented study showed high prediction accuracy. The simplification of the target images may have contributed. The “correctness” of the predictions made by this system may not be guaranteed, but the predictions made in this system were generally satisfactory.

## 1. Introduction

X-ray CT of the craniofacial region is indispensable in obtaining bone information for the diagnosis of craniofacial fractures, facial morphology, and occlusal status. Noise called “metal artifact” appears in the image if there are materials with a high X-ray absorption rate, such as metal prosthetics within the imaging area. Although some efforts(1)(2)(3) are made to reduce these artifacts during imaging, metal artifacts are often seen in the images that clinicians receive. Metal artifacts remain when the bone area is extracted by setting a threshold on CT values, for example, to construct a three-dimensional bony image. The bed of the CT imaging system also remains in the image. It is necessary to manually remove noises from each image using image processing software. These noises disturb observing the 3D models. It is a very time-consuming process. An automatic system was constructed to retrieve bone images without the noises from craniofacial CT images.

## 2. Materials and Methods

All procedures were done on a desk-top personal computer with a GPU: GeForce RTX3090 24.0GB ((nVIDIA, Santa Clara, CA, USA), Windows 10 Pro (Microsoft Corporations, Redmond, WA, USA). Python 3.8 (Python Software Foundation, DE USA): a programing language, was used under Anaconda 15 (FedoraProject. http://fedoraproject.org/wiki/Anaconda#Anaconda_Team_Emeritus) as an installing system, and Spyder 4.1.4 as an integrated development environment. Keras 3 (https://keras.io/): the deep learning library, written in Python was run on TensorFlow 2.5 (Google, Mountain View, CA, USA). GPU computation was employed through CUDA 10.0 (nVIDIA). For 3D reconstruction, slicer 4.11 (www.slicer.org) was used with Jupyter Notebook (https://jupyter.org/). OpenCV 3.1.0 libraries (https://docs.opencv.org/3.1.0/) were used in image processing.

### 2.1 Datasets

#### (1) CT images

From The Cancer Imaging Archive Public Access (wiki.cancerimagingarchive.net), Head-Neck-Radiomics-HN1(4), the collection of CT images from head and neck squamous cell carcinoma patients was retrieved. It consists of the folder of each patient, containing 512×512 pixels DICOM (Digital Imaging and Communications in Medicine) axial images (value ranged 0 to 4071 for each pixel), taken at 5 mm intervals in the cephalocaudal direction. The order of the images was checked and images from the top of the head to the mandible were extracted for 120 cases(5).

#### (2) Bony image segmentation (Fig 1)

**Fig 1.**
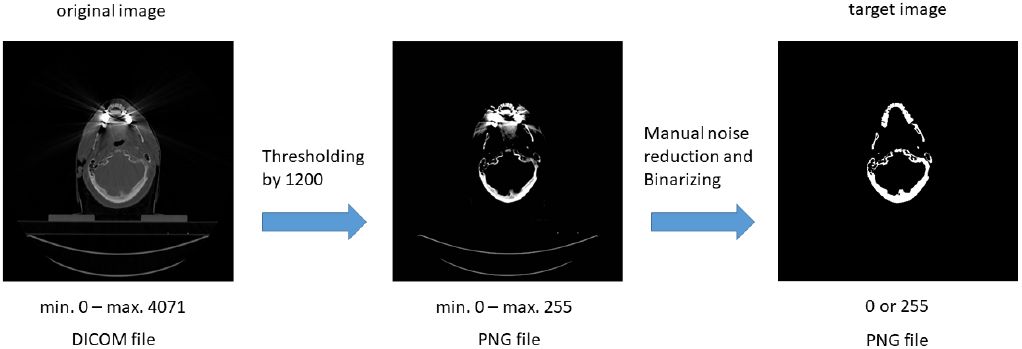
Making target image. The original DICOM images (pixel value ranged 0 to 4071) were thresholded and converted to PNG files. The noises remaining in the files were removed manually. The pixel values were binarized to 0 or 255.

##### *Bone Region Extraction by Threshol*d

Using a Python library: pydicom (https://pydicom.github.io), the slice image was read from each DICOM file. To extract high-density areas, pixel values less than 1200 were replaced with 0. Areas with pixel value greater than 2040 were replaced with 2040. From the thresholded values, 1020 was subtracted and divided by 4. The yield values ranged from 0 to 255. Images were stored as PNG (Portable Network Graphics) files.

##### Manual noise reduction

Noises, such as metal artifacts, beds, etc. in thresholded PNG images were checked one by one visually and removed using image processing software: GIMP (https://www.gimp.org).

##### Binarization

The images were binarized with pixel value 10 as the threshold (0 or 255) and saved as PNG files (target images)

#### (3) Neural network and learning

##### U-net

A U-net model (Fig 2.) was constructed using keras-unet (https://pypi.org/project/keras-unet/). Input and output shapes were 512×512. For final output activation, ReLU (Rectified Linear Unit)(6) was used. Batch normalization option was put on. Dice loss, Jaccard loss and mean squared error were utilized as the loss function respectively. As the optimizer, Adam(7) was used.

**Fig 2.**
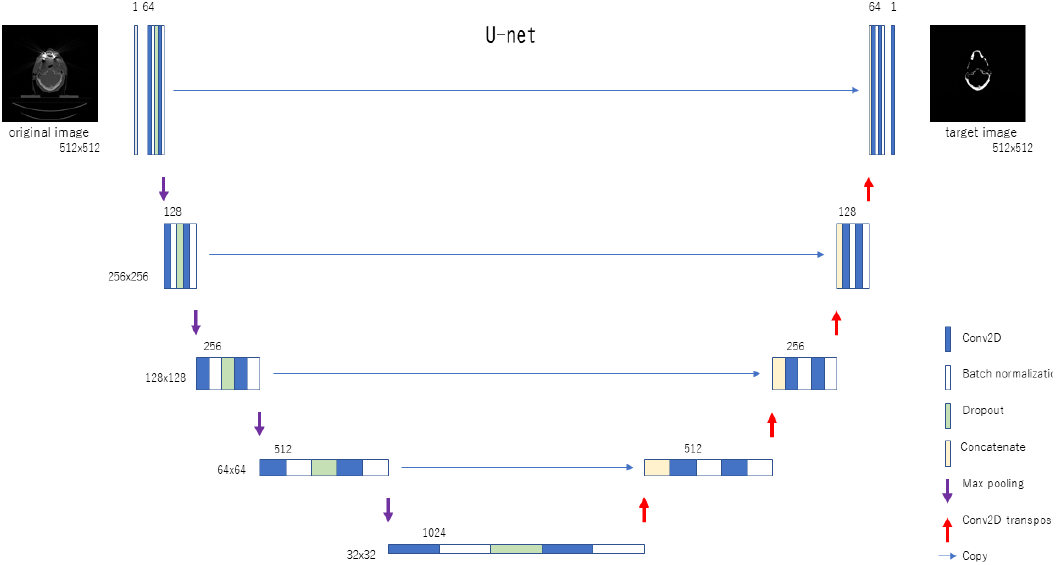
The U-net model used. Input and output were 512 x512.

##### Machine learning

Datasets of the original DICOM image and corresponding target image, counting 7671 pairs, were divided into 5671 training datasets and 2000 validation datasets. DICOM values were divided by 1000 and target image values were divided by 255 to normalize. The U-net models were trained with the training datasets, with early stopping option (https://keras.io/api/callbacks/early_stopping/) and the best weights were saved.

#### (4) Validation

DICOM images of 2000 validation datasets were given to the trained models and predicted images were obtained. The predicted images were binarized by the threshold of 0.5. Mean squared errors between the binarized predicted images and the target images were calculated. Error pixel count for a dataset was obtained by multiplying the mean square error and 512×512.

To visualize, the binarized prediction images were shown in the green channel. The target images were shown in the red channel. When merged, matching pixels were shown yellow. Error pixels were shown green or red.

## 3. Results

Training and validation accuracy with the variety of loss functions is shown in Fig 3. Mean squared errors presented superiority to Dice or Jaccard loss.

**Fig 3.**
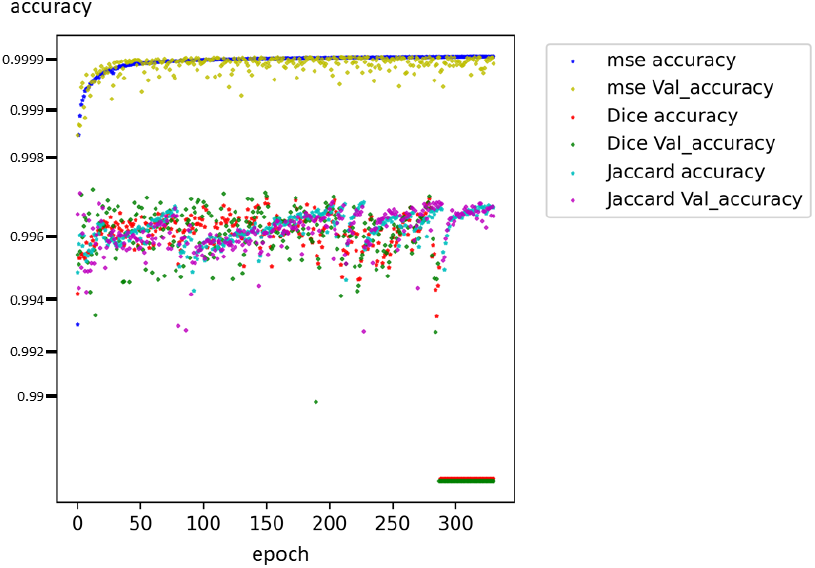
The transition of accuracy with the function loss of mean squared error (mse), Dice loss (Dice), and Jaccard loss (Jaccard)。Vertical axis is shown in logarithm.

The prediction errors and the number of error pixels are shown in Table 1.

**Table 1.**
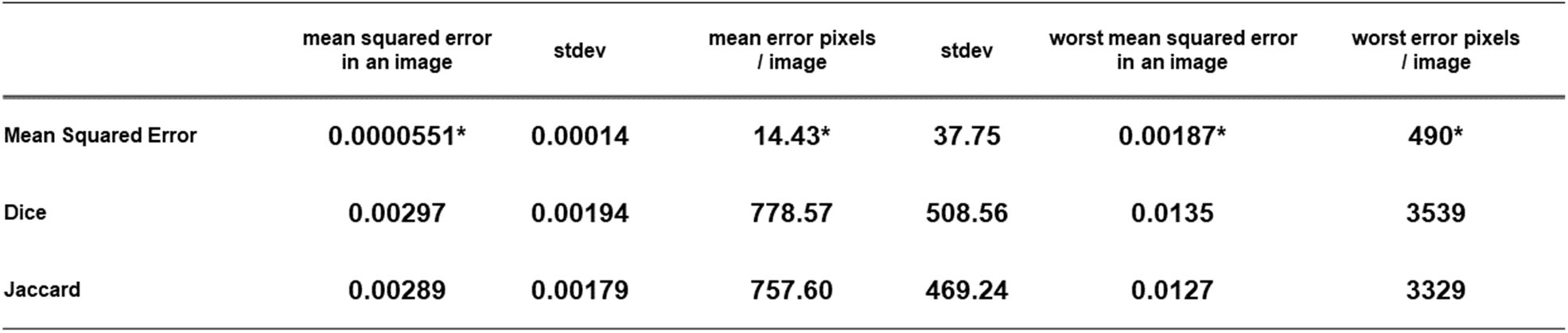
The mean and the worst prediction errors for the 2000 test images, by trained models with different function losses. *: p<0.001 Bonferroni’s test

As an example, predictions of the same image (the worst result with mean squared loss) by the networks trained with the different loss functions are shown in Fig 4.

**Fig 4.**
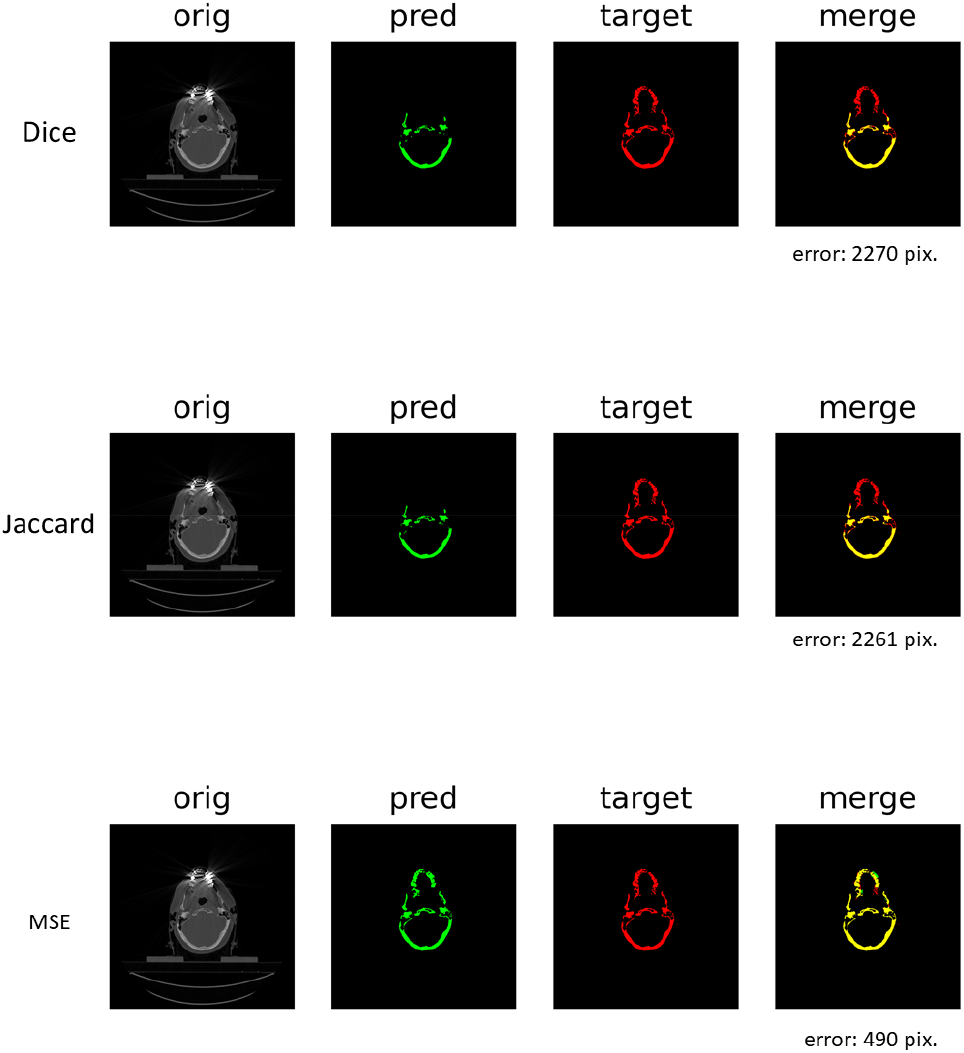
An example of the prediction difference for the same test image (for the model trained with mean squared loss, this image scored the worst). The predicted images are shown in green channel, and the target in red. In the merged image, the correct pixels are shown in yellow or black. The error pixels are shown in green or red.

The merged images with the worst 6 prediction errors by the trained neural networks with each function loss are shown in Fig 5.

**Fig 5.**
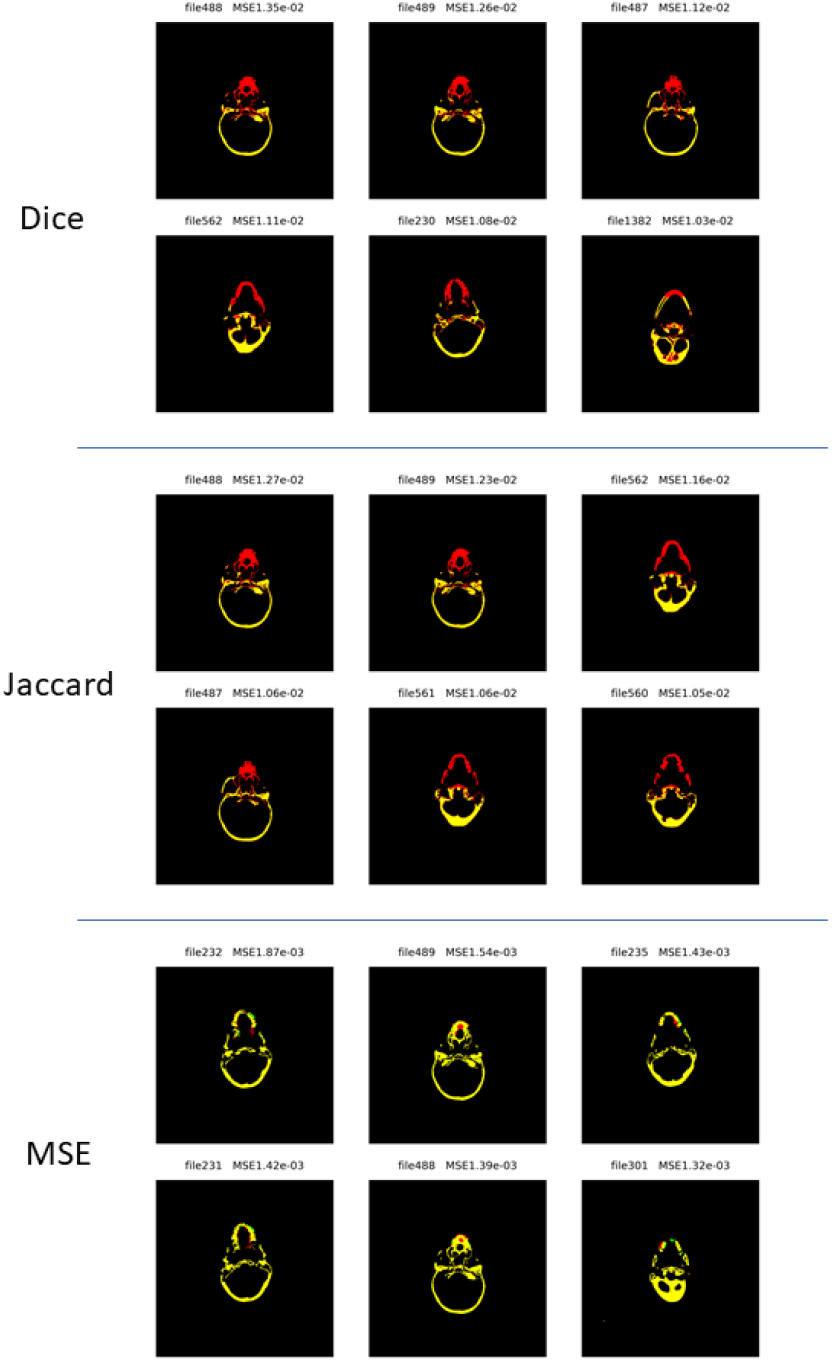
The merged images with the worst 6 errors predicted by the models trained with the three function losses.

A three-dimensional bone reconstruction example of the same original DICOM files, segmented by CT number threshold and neural networks trained with the different loss functions, is shown in Fig6.

**Fig 6.**
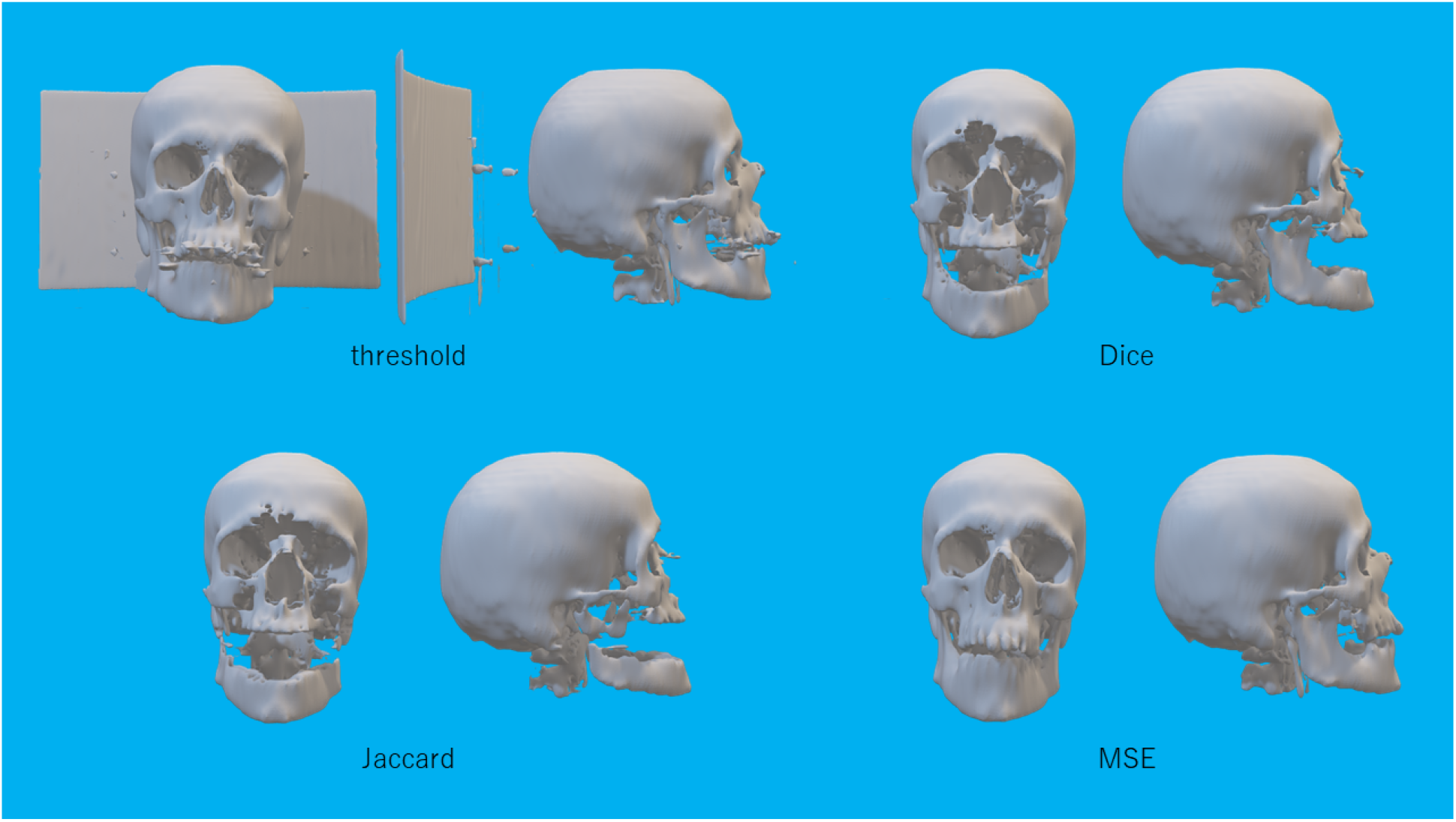
A series of DICOM images for a patient were processed by threshold or predicted by the trained models with three different function losses. The images were over-written on the DICOM files. They were three-dimensionally reconstructed with slicer 4.11.

## 4. Discussion

There have been several reports of metal artifact reduction methods, including utilizing convolution neural networks(8)(9) or generative adverse networks(10)(11). Most of them intended to restore not only the hard tissue but also the soft tissue.

This study is dedicated to the delineation of craniofacial bones. The target images were binarized. The simplification of the target images may have contributed to the high prediction accuracy.

Networks trained with various loss functions showed differences in prediction accuracy. In our setting, mean squared loss presented superiority to Dice loss or Jaccard loss.

The accuracy of machine learning means whether the functional laws hold true in the validation data, that describes the relationship between inputs and outputs in the training data, Even if the prediction accuracy of a well-trained model is high, if the training data itself is not “correct”, the system may not be giving the “correct” answer. In this study, the key to “correctness” was the creation of a bone region image (target images), where artifacts and beds were manually removed. But in the original DICOM images, artifacts mask true information. We had to predict the truth, which is impossible to reproduce completely, with our anatomical and clinical knowledge. Therefore, the “correctness” of the predictions made by this system may not be guaranteed, but we believe that the predictions made by this system are generally satisfactory.

## 5. Conclusion

A U-net based noise reduction system for cranio-facial CT to segment bone was constructed. High prediction accuracy was achieved,

## Data Availability

All data produced in the present study are available upon reasonable request to the authors

## 6. Acknowledgement

There is no financial support.

